# Monitoring trends and differences in COVID-19 case-fatality rates using decomposition methods: Contributions of age structure and age-specific fatality

**DOI:** 10.1101/2020.03.31.20048397

**Authors:** Christian Dudel, Tim Riffe, Enrique Acosta, Alyson van Raalte, Cosmo Strozza, Mikko Myrskylä

**Affiliations:** Max Planck Institute for Demographic Research; Sapienza University of Rome; Interdisciplinary Centre on Population Dynamics (CPop), University of Southern Denmark; Population Research Unit, University of Helsinki

## Abstract

The population-level case-fatality rate (CFR) associated with COVID-19 varies substantially, both across countries time and within countries over time. We analyze the contribution of two key determinants of the variation in the observed CFR: the age-structure of diagnosed infection cases and age-specific case-fatality rates. We use data on diagnosed COVID-19 cases and death counts attributable to COVID-19 by age for China, Germany, Italy, South Korea, Spain, the United States, and New York City. We calculate the CFR for each population at the latest data point and also for Italy over time. We use demographic decomposition to break the difference between CFRs into unique contributions arising from the age-structure of confirmed cases and the age-specific case-fatality. In late April 2020, CFRs varied from 2.2% in South Korea to 13.0% in Italy. The age-structure of detected cases often explains more than two thirds of cross-country variation in the CFR. In Italy, the CFR increased from 4.2% to 13.0% between March 9 and April 22, 2020, and more than 90% of the change was due to increasing age-specific case-fatality rates. The importance of the age-structure of confirmed cases likely reflects several factors, including different testing regimes and differences in transmission trajectories; while increasing age-specific case-fatality rates in Italy could indicate other factors, such as the worsening health outcomes of those infected with COVID-19. Our findings lend support to recommendations for data to be disaggregated by age, and potentially other variables, to facilitate a better understanding of population-level differences in CFRs. They also show the need for well designed seroprevalence studies to ascertain the extent to which differences in testing regimes drive differences in the age-structure of detected cases.

## Introduction

The novel Coronavirus disease 2019 (COVID-19), caused by severe acute respiratory syndrome coronavirus 2 (SARS-CoV-2), has been spreading rapidly across the world, and on March 11 2020 was recognized as a pandemic by the World Health Organization.

COVID-19 outbreaks went along with mostly regular patterns of logarithmic increase of the number of confirmed cases, with a few notable exceptions. The number of deaths associated with COVID-19, however, have evolved considerably less regularly, and case-fatality rates (CFRs) differ substantially between countries [1,2].

Examples of this discrepancy are shown in Fig 1. As of April 22, 2020, Germany had a total of around 150 thousand confirmed infections and 5700 deaths, resulting in a CFR of around 3.8%. Italy, on the other hand, up to the same day, had close to 174 thousand confirmed cases of infection, around 23 thousand deaths, and a CFR of 13.0%. On April 13, Italy had roughly the same number of cases as Germany on April 22, and a CFR of 12.7%. Thus, the outbreak in Italy is going along with a much higher CFR, which has also increased over time [2,3].

**Fig 1.**
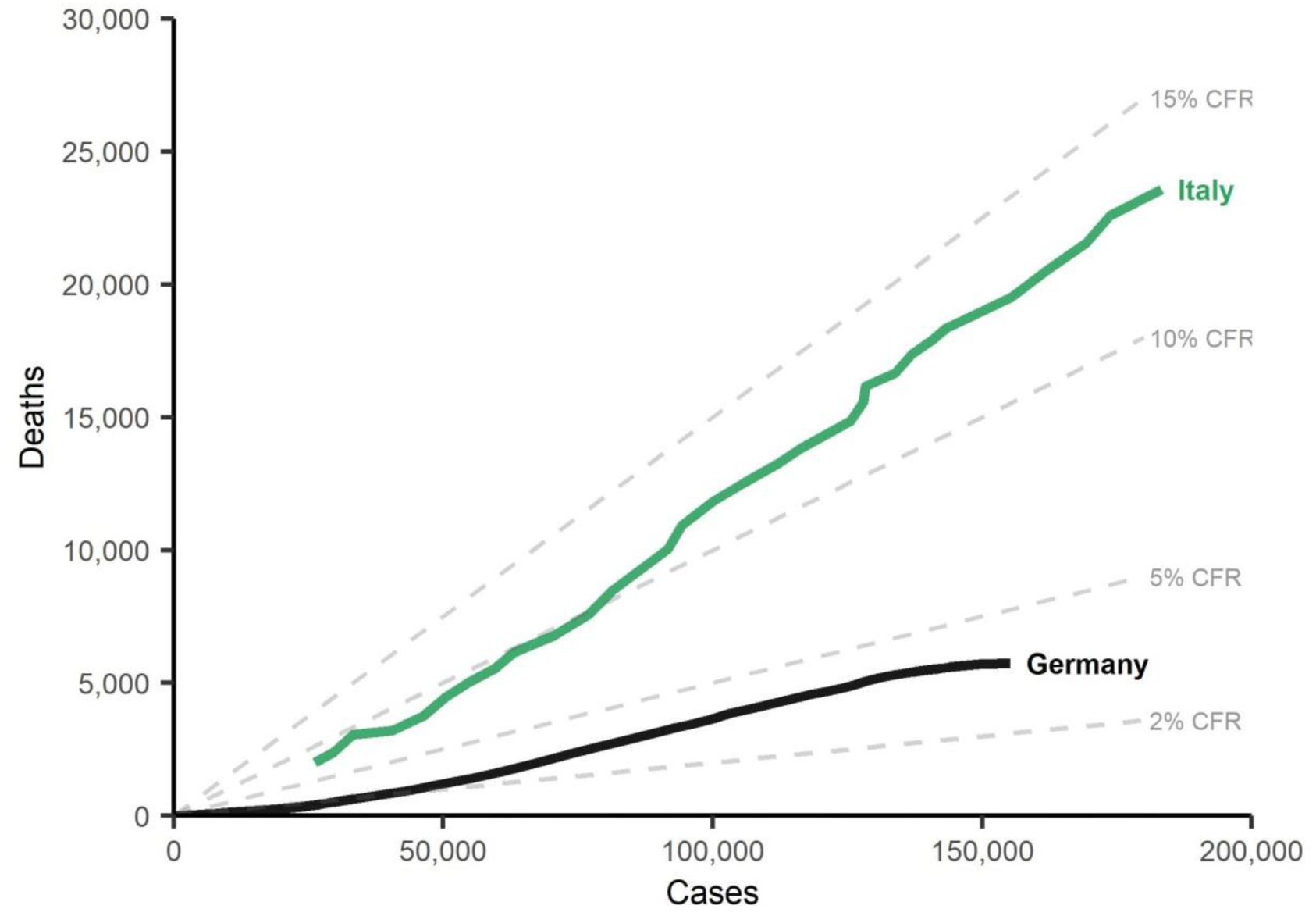
COVID-19 confirmed cases and deaths, and implied case-fatality rates (CFR) in Italy and Germany since March 1, 2020 (Germany) and March 9, 2020 (Italy).

Differences in the CFR could indicate that the risk of dying of COVID-19 among detected cases differs between countries or changes within a population over time. On the other hand, it could also imply compositional differences in the detected infections [1,3]. Specifically, the risk of dying of COVID-19 is well-documented to increase with age. Thus, if the population of confirmed infected individuals is older in one country or time period than in another, the CFR will be higher, even if the age-specific risk of dying is the same.

Indeed, demographers have argued that age structure matters, and the age composition of the reported cases has been suggested as a potential explanation for differences in CFRs [1-5]. So far, however, there have been no assessments of the importance of the age structure of diagnosed cases versus the age-specific CFR.

In this paper, we analyze cross-country differences in observed CFRs and within-country time trends in CFRs. We use recent data on China, Germany, Italy, South Korea, Spain, the United States, and New York City. We use a standard demographic decomposition technique to disentangle two potential drivers of differences and trends: (1) the age structure of confirmed cases and (2) age-specific case-fatality rates [6].

We interpret our findings in light of the unfolding knowledge about data-driven biases affecting CFRs. Counts of confirmed cases and deaths might not be comparable across countries because of differences in case and death definitions; differences in the underestimation of cases and in their age structure as a consequence of the country-specific testing regime; reporting delays of case counts and death counts; and differences in delays between symptoms and death [1,2]. These data-related issues might lead to over- or under-estimation of CFRs throughout the epidemic, and more reliable estimates will only be available after its conclusion. Currently, adjusting CFRs for all of these potential biases is challenging and beyond the scope of this paper. Nevertheless, the method described in this paper is also readily applicable to adjusted estimates of CFRs once they become available.

Decomposition approaches like the one used in this paper are commonly used to explain the role of age structure on changing incidence rates [7]. They have also been applied to differences in cancer fatality rates across regions with varying age structures [8]. We are not aware of any application to CFRs of infectious diseases in general and the COVID-19 pandemic in particular.

To facilitate the application of the approach described in this paper, we provide code and reproducibility materials for the open source statistical software R in a freely-accessible repository on the Open Science Framework: https://osf.io/vdgwt/. Moreover, we also provide some examples in an Excel spreadsheet in the same repository, and an interactive spreadsheet on Google Docs: https://bit.ly/2WVkPAg.

## Data and methods

### Data

We gathered data on the cumulative number of diagnosed infections and deaths attributable to COVID-19 for the following populations (in alphabetical order): China, Germany, Italy, South Korea, Spain, the United States, and New York City. An overview of the data is given in Table 1. For Italy, we include a total of 8 days in the analysis, covering the period starting on March 9 and ending on April 22. For all other populations, we use the data of the most recent date. For China, the most recent available age-specific data is from February, while for the remaining cases the data is from April 22.

**Table 1:** Populations covered in the analysis, and their cumulative detected cases and deaths. For most, we use the latest data available to us. The age-specific data for China does not account for the retrospective correction of the number of deaths. For Italy, we also analyze trends over time. The cumulative cases and deaths shown for Italy in this table are for April 22.

| **Country** | **Date(s)** | **Detected cases**<br>**(cumulative)** | **Deaths**<br>**(cumulative)** |
| --- | --- | --- | --- |
| China | February 11 2020 | 44,672 | 1,017 |
| Germany | April 22 2020 | 149,993 | 5,711 |
| Italy | March 9, March 16-April 22 2020 | 173,730 | 22,586 |
| South Korea | April 22 2020 | 10,649 | 238 |
| Spain | April 22 2020 | 213,024 | 22,157 |
| United States | April 22 2020 | 828,441 | 46,379 |
| New York City (US) | April 22 2020 | 141,754 | 10,290 |

All data is provided by the respective health authorities, and is collected as part of another project which gathers and standardizes age-specific data on the COVID-19 pandemic [9]. The database is continuously updated and freely available online, but we also provide snapshots of the data used for the calculations in this paper together with the code. A complete list of sources is provided in the documentation of the database project [9].

For some of the countries (Germany, Italy, Spain, United States, and New York City) age is not available for some confirmed cases or deaths. The database project imputed the missing age using the observed age distribution of cases or deaths, respectively [9]. Removing these cases from the analysis altogether has no substantive impact on the results, except for Spain, where around 28% of cases and 44% of deaths have no recorded age. Ignoring cases and deaths of unknown age in Spain would therefore deflate age-specific case-fatality rates.

The original data is provided in different age groupings. For the decomposition, the age groups have to match. The database project adjusted counts so that all countries conform with the age groups of South Korea, for which the age groups are 10-year age groups from birth to 80+. Counts were split using a recently proposed method tailored for this data situation [9,10]. The supplementary materials show the original age groups of the data.

### Case-fatality rates

The COVID-19 case-fatality rate (CFR) is defined as the ratio of deaths (D) associated with COVID-19 divided by the number of detected COVID-19 cases (N): CFR=D/N. In our application, the death and case counts are cumulative counts up to a certain date.

If case counts and death counts are available by age, which is our situation, the CFR can also be written as a sum of age-specific CFRs weighted by the proportion of cases in a certain age group. We use *a* as an index to denote different age groups. These age groups could, for instance, be 0 to 9 years, 10 to 19 years, and so on, but other groupings are also possible. We define age-specific CFRs as 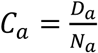; i.e., the number of deaths in age group *a* divided by the number of cases in the same age group. The proportion of cases in age group *a* is given by 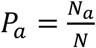. Using this notation, the CFR can be written as a weighted average of age-specific CFRs:

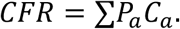

We use the weighted expression and a mathematical decomposition approach introduced by Kitagawa to separate the difference between two CFRs into two distinct parts, one attributable to age-structure and another to age-specific case-fatality [6]. The method attributes the total difference into these two components, leaving no residual. In other words, if we use *i* and *j* to index two different populations, then the decomposition approach splits the difference between their CFRs into

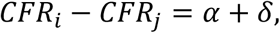

where the *α*-component captures the effect of the age structure, and the *δ*-component indicates the part of the difference attributable to age-specific case-fatality. The details of the method are described in the supplementary materials, which also provides a step-by-step walk-through of the decomposition.

## Results

### Country comparisons

Table 2 shows results for cross-country comparisons using the data from South Korea (April 22) as a reference, with countries sorted by increasing CFR. We chose South Korea as the reference because its CFR is arguably the closest match to its actual infection rate due to extensive testing and an earlier onset of the epidemic; moreover, the CFR was comparably low, and decompositions will estimate what factor leads other countries to differ from this low CFR setting, making results easy to interpret. For all other countries, we use the most recent date available to us, as shown in Table 1. In the supplementary materials, we provide additional results using Germany (low CFR) and Italy (high CFR) as reference countries.

**Table 2:** Results of the cross-country decompositions of case-fatality rates (CFRs) using South Korea as a reference case. The third column shows the difference between each country and South Korea as a reference, and is calculated as the CFR of South Korea minus the CFR of the respective country. Data for all countries is for April 22, except China (February 11).

| Country | CFR | Difference | Age ($\alpha$) component | Fatality ($\delta$) component | Age ($\alpha$) component, relative | Fatality ($\delta$) component, relative |
| --- | --- | --- | --- | --- | --- | --- |
| (1) | (2) | (3) | (4) | (5) | (6) | (7) |
| South Korea | 0.022 |  |  | (Reference) |  |  |
| China | 0.023 | -0.001 | -0.002 | 0.001 | 60.1% | 39.9% |
| Germany | 0.038 | -0.016 | -0.014 | -0.002 | 88.0% | 12.0% |
| USA | 0.056 | -0.034 | -0.014 | -0.019 | 42.9% | 57.1% |
| New York City | 0.073 | -0.050 | -0.013 | -0.037 | 26.3% | 73.7% |
| Spain | 0.104 | -0.082 | -0.057 | -0.025 | 69.2% | 30.8% |
| Italy | 0.130 | -0.108 | -0.071 | -0.037 | 65.7% | 34.3% |

Based on the cumulative data up to April 22, South Korea had a CFR of 2.2% (first line of the table, column CFR). For all countries the difference to the South Korean CFR is shown in the third column of the table (South Korea minus the respective country). The fourth and fifth column of the table show the absolute contributions of the case age distribution and age-specific fatality components, respectively. A negative number for the age structure indicates an older age structure of detected cases compared to South Korea, while a negative number for the fatality component indicates higher age-specific case-fatality rates compared to South Korea. The sixth and seventh column of the table indicate the relative contributions of the components.

All countries and regions have a higher CFR than South Korea, as indicated by the negative difference shown in column four of Table 2, and some of the differences are substantial. For instance, the Italian is almost six times as high.

In many cases, the (relative) contributions of the *α*-component (age structure) seem to be larger than the *δ*-component (fatality), and the *α*-component is always negative. This means that the age structure of cases is an important factor in explaining why most countries we study fare worse than South Korea. For instance, in the two cases with the highest CFRs – Italy and Spain – the relative contributions were similar with the *α*-component explaining around two thirds of the difference (Italy: 65.7%; Spain: 69.2%), and the *δ*-component explaining the remainder. In Germany, the case age structure is the main driver of the difference in CFRs relative to South Korea, and explains close to 90% of the difference. The US and New York City seem to be an exception, and the high CFR compared to South Korea seems to be mostly due to higher mortality of infected individuals.

### Trends over time: The case of Italy

For Italy we have a relatively long time series of data spanning several weeks. Table 3 documents how the Italian CFR evolved from March 9 to April 22, with selected dates presented in between. The CFR of March 9 is used as a reference, and the decomposition shows which factor is driving the trend in the CFR. From the beginning to the end of the period under study the CFR almost tripled, from 4.3% to 13.0%. This increase over time is largely driven by worsening fatality of COVID-19 – the fatality component explaining more than 80% of the rise in almost all time periods – and changes in the age structure of cases only played a minor role, with detected cases moving to a more favorable (younger) age distribution and slightly counteracting the effect of worsening fatality. As a robustness check we changed the reference period from March 9 to March 21 (CFR: 8.1%). This resulted in the fatality component explaining more than 90% of the increase in CFR.

**Table 3:** Development of the Italian case-fatality rate (CFR) over time. The third column gives the difference between the CFR of the respective date minus the CFR of March 9.

| Date<br>(1) | CFR<br>(2) | Difference<br>(3) | Age ($\alpha$) | Fatality ($\delta$) | Age ($\alpha$) | Fatality ($\delta$) |
| --- | --- | --- | --- | --- | --- | --- |
|  |  |  | component<br>(4) | component<br>(5) | component,<br>relative<br>(6) | component,<br>relative<br>(7) |
| 09 March 2020 | 0.043 |  | (Reference) |  |  |  |
| 16 March 2020 | 0.068 | 0.025 | -0.010 | 0.035 | 22.84% | 77.16% |
| 22 March 2020 | 0.088 | 0.046 | -0.009 | 0.054 | 13.87% | 86.13% |
| 28 March 2020 | 0.104 | 0.061 | -0.009 | 0.070 | 10.89% | 89.11% |
| 3 April 2020 | 0.118 | 0.075 | -0.012 | 0.087 | 12.05% | 87.95% |
| 9 April 2020 | 0.122 | 0.080 | -0.012 | 0.091 | 11.28% | 88.72% |
| 15 April 2020 | 0.125 | 0.083 | -0.010 | 0.093 | 9.80% | 90.20% |
| 22 April 2020 | 0.130 | 0.087 | -0.008 | 0.095 | 7.82% | 92.18% |

## Discussion

Case-fatality rates (CFRs) associated with COVID-19 vary strongly across countries and over time within countries. Our findings show that there is substantial variation in which factor explains the differences in CFRs. Differences in the age distribution of detected infections in some cases explain a substantial part of the total difference in CFRs. In particular, more than 50% of the difference in CFRs between countries with a low CFR and a high CFR can be explained by the age structure of detected infections. In contrast, in Italy, we observe a substantial increase in the CFR over time, mostly attributable to increasing age-specific case-fatality.

Ultimately, the approach discussed here does not directly explain why the age structure of confirmed cases or the age-specific case-fatality rates matter more in one case and less in another, and some expertise about the contexts which are being compared is required to interpret results. We discuss some potential explanations below, including potential data-related issues and biases.

Differences in the age structure of the populations which are being compared are unlikely to be a major driver of the age component that we estimated here, as the age composition of confirmed cases does not necessarily match the age composition of the population. For instance, according to Eurostat, the proportion of the population aged 80+ in 2019 was 7% in Italy and 6.5% in Germany, while in our data the proportion of reported infections in the same age range was 24% for Italy and only 11% in Germany.

Differences in testing regimes are a plausible mechanism driving both the different age structures of detected cases, as well as different age-specific fatality rates to the extent that denominators are underestimated in distinct degrees [3,11,12]. Results not shown here indicate that early in the pandemic in March the difference in the CFRs of South Korea and Germany - two countries with extensive and early tracing and testing of contacts of known cases - was largely driven by differences in fatality. The low contribution of the case age distribution component to the CFR disparity between South Korea and Germany suggests that these countries might have been more successful at catching the mild and asymptomatic cases among the younger population groups. Since then, the CFR of Germany has increased and the age structure of confirmed cases has shifted to higher ages, and the age structure has become more important in explaining the gap between South Korea and Germany, making test numbers alone an unlikely explanation for the different age structure of detected cases.

Differences in the COVID-19 transmission pathways might also be a factor. Depending on contact patterns and household structure, the elderly population might be affected earlier in some countries than in others, leading to a less favorable age distribution of infections [4,13]. This could be relevant in explaining why the age distribution plays such a large role for the two countries with by far the highest CFR, Spain and Italy, which have a relatively large proportion of individuals living with their elderly parents or grandparents, and comparatively intensive intergenerational contact [14-16].

Disparities in age-specific case-fatality rates across countries may result from either differences in age-specific prevalence of comorbidities, which exacerbate the risk of death from COVID-19 considerably [1,17] or differences in quality or saturation levels of the healthcare system. The trend over time in the Italian CFR is an example where changes in age-specific case-fatality rates are driving trends, instead of changes in the case age distribution. This likely reflects the worsening situation in Italy over time as its health care system got under increasing pressure [11,18]. However, an increase in CFR could also be expected once containment measures become effective, and newly confirmed cases increase at a slower pace than deaths from cases acquired prior to containment policies.

Only once an epidemic reaches its final conclusion and all cases have either resulted in recovery or fatalities, can the importance of the age difference in cases on CFRs be assessed with an acceptable degree of accuracy [19]. In this context a distinction should be made between CFRs, which are solely based on detected cases, and infection fatality rates (IFRs), which estimate the risk of dying once infected, including both confirmed and undiagnosed cases. Ideally policies for containing the spread of a virus would be designed on the basis of IFRs. However, particularly early on in an epidemic, the CFR is the only metric available until the extent of known data-driven biases can be assessed [1,11,12,20-23].

Data quality can affect both the age composition of detected cases and age-specific case-fatality rates. For instance, counts may be affected by issues like reporting delays or censoring, or by inconsistent case definitions [1,2,20,21,24]. In many countries, only deaths occurring in hospitals are being reported in a timely manner [25], underestimating the full death count which would include deaths at home and in institutions. Deaths may be underestimated because of a lack of testing both before and after death. Countries might also differ in how they code deaths from underlying or contributory causes [25]. Excess all-cause mortality compared to a seasonal allcause mortality baseline are suggestive that there is currently considerable underreporting of COVID-19 deaths, even if some of these deaths might be related to delayed or avoided medical treatment from other causes of death [26].

The relative importance of both the case age structure and mortality components could also be affected by comparing countries at different stages of the epidemic. This could result from cases not being detected at the beginning of the epidemic, or from differences in the lag between infection and death [11,23,27]. Generally, CFRs are highest at the beginning of an infectious outbreak, when the most serious cases are the most readily detected, and declines as testing capacity increases and less serious cases are identified [23]. This has notably not been the case for the COVID-19 epidemic, where the CFR has generally been increasing. Likely this reflects the success of widespread containment measures enacted in response to increasing caseloads. Newly identified cases are increasing more slowly than deaths, despite increases in testing capacity.

The application of the method we present in this paper is not limited to decomposing the current estimates of CFRs. It can also be applied to CFR estimates which have been corrected for biases, to infection fatality rates (IFRs) - which estimate the risk of dying from both detected and undetected cases -, and to excess all-cause weekly mortality counts. Thus, while the data currently available as input for the decomposition approach might be of varying quality, this is not a flaw of the method itself. As data quality improves over time and adjustment methods become available our approach will continue to provide insights into differences and trends in mortality associated with COVID-19.

Finally, the choice of age groups may have affected our results. If ages were grouped too widely it might hide actual age-specific case-fatality differences. For instance, if the median age within the 10-year aggregated age groups that we used differed between populations, this would reduce the case-age structure explanation and inflate the age-specific mortality explanation. Finally, there are alternative decomposition techniques that might yield different results. However, differences are expected to be rather small; indeed, applying the method of Horiuchi and colleagues [28] to our data yields virtually the same results (results available upon request).

The results of this study add weight to recommendations for data to be disaggregated by age and potentially other variables to facilitate a better understanding of population-level differences in CFRs. Equally important will be well designed seroprevalence studies to ascertain the extent to which our findings are driven by differences in testing regimes, particularly in the diagnosis of mild and asymptomatic cases. To this extent we are encouraged by the recent announcement that such a study is being initiated in Germany in line with official WHO recommendations [29,30].

Overall, our results show that differences between countries with low and high CFRs can be driven to a significant extent by the age structure of confirmed cases. Decomposing differences in case-fatality rates over time or between countries reveals important insights for monitoring the spread of COVID-19. An accurate assessment of these differences in CFR across countries and over time are crucial to inform and determine appropriate containment and mitigation interventions, such as social confinement and mobility restrictions.

## Data Availability

All data and code is available online.

https://osf.io/vdgwt/

## Acknowledgements

We thank Catalina Torres for collecting parts of the data and for helpful comments and suggestions.

## A. Details of the decomposition method

### A.1 Decomposing CFRs

We want to decompose, or “explain”, the difference between two CFRs, irrespective of whether they are from two different populations, or from the same population at two different points in time, or from different groups within a population, e.g., gender or socio-economic groups. We will use *CFR_i_* and *CFR_j_* to distinguish the two CFRs, e.g., country *i* and country *j*. Moreover, we write *P_ia_*, *C_ia_*, *P_ja_*, and *C_ja_* for the underlying age compositions and age-specific CFRs; i.e., *CFR_i_ = ΣP_ia_C_ia_*.

Using a decomposition approach introduced by Kitagawa [6] we separate the difference between two CFRs into two distinct parts,

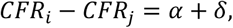

where *α* captures the part of the difference between CFRs which is due to differences in the age composition of cases, and *δ* is due to differences in mortality. *α* is given by

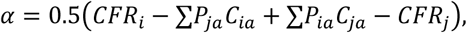

while *δ* can be calculated as

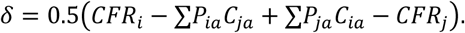

Note that the age groups for group *i* and *j* group need to be the same. If this is not the case in the raw data and, for instance, one country reports counts in 5-year age groups (0-4, 5-9, 10-14, 15-19, …) and the other uses 10-year age groups (0-9, 10-19, …), then either the more finely grained data needs to be aggregated to match the coarser data, or the coarser data needs to be adjusted. We choose the latter approach (see appendix C below).

The intuition behind the formulas is as follows. The first two terms in brackets in the equation for *α* are *CFR_i_ – ΣP_ja_C_ia_*, or, replacing *CFR_i_* with its definition, *CFR_i_ – ΣP_ja_C_ia_*. The second sum in this expression captures how high the CFR would have been if group *i* had the same age distribution of infections as group *j*. The difference to the actual CFR (the whole expression) then captures to what extent the CFR is higher than this hypothetical CFR because of the actually observed age distribution of detected infections. The third and the fourth term in brackets in the equation for *α* are following a similar logic, but using a different hypothetical comparison, asking how much the CFR of group *j* would differ if the detected cases had the age distribution of group *i*. The formula for *δ* again follows a similar logic, but now replacing the age-specific CFRs instead of the age distribution. In summary, to decompose the difference between two CFRs requires nothing more than the two CFRs themselves as well as a few additional hypothetical CFRs.

To calculate the proportion *α* and *δ* contribute to the total difference one can use 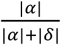 in case of *α* and 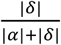 for the contribution of *δ*.

### A.2 Interpretation

As an artificial example, assume that the CFR in country A is equal to 2 percent, while it equals 4 percent in country B. Subtracting the CFR of country A from country B gives a difference of 2 percentage points. If a large part of this difference is due to the age structure, then *α* could be 0.015 and *δ* could be 0.005. These sum to 0.02, or 2 percentage points. If, as another example, two countries have the same age structure of cases, then *α* will be zero. A similar reasoning holds for *δ* if age-specific CFRs are the same for both countries being compared. In relative terms, the *α*-component explains 75 percent of the difference between countries, while the *δ*-component only explains 25 percent.

The total difference between two CFRs as well as both *α* and *δ* can be negative. The formulas for the relative contributions take this into account by using absolute values. If the total difference is positive and either *α* or *δ* are negative, it means that the corresponding part of the difference actually reduces the difference between CFRs. For instance, when comparing the CFR for one country at two points in time, the total difference could be 0.03; i.e., the CFR increased by three percentage points. If in this case *α* would be negative, say −0.01, it would mean that the age distribution of cases over time became more favorable. *δ* would be 0.04 in this scenario, and without changes in the age distribution of infections as captured through *α*, the difference between CFRs would even have increased by four percentage points.

## B. Additional results

**Table 1:** Results of the cross-country decompositions using Germany as a reference case.

| Country<br>(1) | CFR<br>(2) | Difference<br>(3) | Age ($\alpha$)<br>component<br>(4) | Fatality ($\delta$)<br>component<br>(5) | Age ($\alpha$)<br>component,<br>relative<br>(6) | Fatality ($\delta$)<br>component,<br>relative<br>(7) |
| --- | --- | --- | --- | --- | --- | --- |
| South Korea | 0.022 | 0.016 | 0.014 | 0.002 | 88.0% | 12.0% |
| China | 0.023 | 0.015 | 0.008 | 0.007 | 55.5% | 44.5% |
| Germany | 0.038 |  |  | (Reference) |  |  |
| USA | 0.056 | -0.018 | 0.000 | -0.018 | 0.9% | 99.1% |
| New York City | 0.073 | -0.035 | 0.008 | -0.043 | 16.1% | 83.9% |
| Spain | 0.104 | -0.066 | -0.035 | -0.031 | 53.1% | 46.9% |
| Italy | 0.130 | -0.092 | -0.049 | -0.043 | 52.9% | 47.1% |

**Table 2:** Results of the cross-country decompositions using Italy as a reference case

| Country<br>(1) | CFR<br>(2) | Difference<br>(3) | Age ($\alpha$)<br>component<br>(4) | Fatality ($\delta$)<br>component<br>(5) | Age ($\alpha$)<br>component,<br>relative<br>(6) | Fatality ($\delta$)<br>component,<br>relative<br>(7) |
| --- | --- | --- | --- | --- | --- | --- |
| South Korea | 0.022 | 0.108 | 0.071 | 0.037 | 65.7% | 34.3% |
| China | 0.023 | 0.107 | 0.053 | 0.054 | 49.4% | 50.6% |
| Germany | 0.038 | 0.092 | 0.049 | 0.043 | 52.9% | 47.1% |
| USA | 0.056 | 0.074 | 0.066 | 0.008 | 89.0% | 11.0% |
| New York City | 0.073 | 0.057 | 0.067 | -0.010 | 87.1% | 12.9% |
| Spain | 0.104 | 0.026 | 0.013 | 0.013 | 48.4% | 51.6% |
| Italy | 0.130 |  |  | (Reference) |  |  |

## C. Additional information on data: Age groups

The data we use is provided in different age groups, depending on the country. The following age groups are used in the original data for both case counts and death counts:

- China: 0-9, 10-19, 20-29, 30-39, 40-49, 50-59, 69-69, 70-79, 80+
- Germany: 0-4, 5-14, 15-34, 35-59, 60-79, 80+
- Italy: 0-9, 10-19, 20-29, 30-39, 40-49, 50-59, 69-69, 70-79, 80-89, 90+
- Spain: 0-9, 10-19, 20-29, 30-39, 40-49, 50-59, 69-69, 70-79, 80+
- South Korea: 0-9, 10-19, 20-29, 30-39, 40-49, 50-59, 69-69, 70-79, 80+
- United States: 0-19, 20-44, 45-54, 55-64, 65-74, 75-84, 85+
- United States (New York City): 0-18, 18-44, 45-64, 65-74, 75+

For the decomposition, the age groups have to match. This is the case for China, Spain, South Korea, and Italy; in the case of the latter the age categories 80-89 and 90+ have to be merged. The age groups provided for Germany and the United States are problematic, as they do not match the age groups of any other country. Aggregating the age groups, as for Italy, does not help, either. For instance, the age category of 20 years to 44 years available for the US cannot be created based on the German data. To deal with this issue, the database project from which we obtained the case and death counts [9] uses a smoothing approach to estimate counts for age groups 0-9, 10-19, …, 80+ [10].

## D. Excel spreadsheet and R code

All R code used to produce the results in the paper as well as the data can be downloaded from https://osf.io/vdgwt/. An Excel spreadsheet containing several examples can be found in the same repository. An interactive spreadsheet is available on Google Docs: https://bit.ly/2WVkPAg.

